# A modified SEIR meta-population transmission based Modeling and Forecasting of the COVID-19 pandemic in Pakistan

**DOI:** 10.1101/2020.06.03.20121517

**Authors:** Sohaib Hassan, Bilal Javed Mughal, Marian Siwiak, Zafar Yasin

## Abstract

The coronavirus disease 2019 (COVID-19) started from China at the end of 2019, has now spread across the globe. Modeling and simulation of the COVID-19 outspread is significant for timely and effective measures to be taken. Scientists around the world are using various epidemiological models to help policymakers to plan and determine what interventions and resources will be needed in case of a surge and to estimate the potential future burden on health care system. Pakistan is also among the affected countries with 18^th^ highest number of total detected number of cases, as of 3^rd^ of June, 2020. A modified time-dependent Susceptible-Exposed-Infected-Recovered (SEIR) metapopulation transmission model is used in the Global Epidemic and Mobility Model (GLEaM) for this simulation. The simulation assumes the index case in Wuhan, China and models the global spread of SARS-COV-2 with reasonable results for several countries within the 95% confidence interval. This model was then tuned with parameters for Pakistan to predict the outspread of COVID-19 in Pakistan. The impact of Non-Drug Interventions on “flattening the curve” are also incorporated in the simulation and the results are further extended to find the peak of the pandemic and future predictions. It has been observed that in the current scenario, the epidemic trend of COVID-19 spread in Pakistan would attain a peak in the second decade of month of June with approximately (3600-4200) daily cases. The current wave of SARS-COV-2 in Pakistan with is estimated to cause some (210,000 – 226,000) cumulative cases and (4400-4750) cumulative lost lives by the end of August when the epidemic is reduced by 99%. However, the disease is controllable in the likely future if inclusive and strict control measures are taken.

## 1 Introduction

COVID-19 started from city of Wuhan, China in December 2019 has spread in all countries becoming a global challenge. The disease caused by corona virus has also been characterized as a pandemic by WHO [1]. Public health efforts to control the COVID-19 heavily depend on how this spreads across the globe. Until today 6484853 confirmed cases and 383816 deaths (as of 3^rd^ of June, 2020) of COVID-19 have been announced in the whole world [2]. US is on the top COVID-19 effected countries and Pakistan is the 18^th^ with 80, 788 cases and 1632 deaths (as of 3^rd^ of June, 2020). The Pakistani government has implemented severe measures to mitigate the outbreak.

The fatality rate for a pandemic depends on many factors including how many people got infected, how the virus spreads and there is not a single fatality rate as it also depends on age along with other many factors and reports vary from country to country. Due to many factors, there could also be inaccuracy in available data. Ideally everyone having signs of infection of the coronavirus should be tested, as well random screening tests shall be performed. This is necessary to know the real number of infections and deaths. Furthermore, transmission is not going to be the same from one country to another due to local conditions and political decisions, etc. That is why some modeling parameters could vary from one country to another.

Modeling and simulations have a major role in scientific research and are being widely used in every field of science and technology including prediction of epidemics. Mathematical models predict the cause of spread of epidemic and provide guidelines for its control. These computational models are developed using well known equations. Computational modeling of COVID-19 spread can help to defeat the virus and plays a vital role in understanding the outbreaks of epidemics and provide valuable material to decision makers to revise their strategy to take precautionary measures timely and to make standard operated procedures(SOPs).

In the present study, the modeling of COVID-19 pandemic in Pakistan is performed using the GLEAMviz simulator framework [3-5]. This package was already used successfully in the past to predict spreading of A/H1N1 and the accuracy of the simulated results was later confirmed [6]. The SIR (Susceptible-Infectious-Removed) model is used where individuals infect each other directly. Susceptible-Exposed-Infectious-Removed (SEIR) model is a modified form of SIR model. In the present work, we have used a modified form of SEIR metapopulation transmission model based on study of global spread of COVID-19 [1].

We compared our results with the actual number of diagnosed cases and found to be in good agreement. Model predictions indicate the peak in Pakistan would arrive in the second decade of June, 2020 with (3600 – 4200) daily cases and start decreasing from there with rate depending upon the interventions by authorities. However, these predicted values strongly depend on the social distancing measures adopted by the public, diagnostic facilities and government’ public health interventions. The current wave of SARS-COV-2 in Pakistan with is estimated to cause some (210,000 – 226,000) cumulative cases and (4400-4750) cumulative lost lives by the end of August when the epidemic is reduced by 99%. However, the disease is controllable in the likely future if inclusive and strict control measures are taken.

## 2 Modeling using GLEAMviz

The SEIR is a basic model used for modeling the epidemics. The SEIR model divides the whole population into four categories: susceptible-the people who could potentially catch the disease; the exposed-people who might have the virus but show no symptoms; the infectious-active cases; and the recovered.

For modeling and simulation, the GLEAMviz is used. It is designed for epidemiology and could be used for modeling the outbreak of epidemics on regional and across the globe. Recently it has already been used for COVID-19 spread prediction [1,7]. It uses the stochastic computational model that integrates demographic and mobility data and uses a compartmental approach to define the epidemic characteristics of the infected disease. Model also takes into account the population and mobility data. Here we have used the version 7 of the GLEAMviz. We have used a modified SEIR model in the present modeling [1]. More details on the framework can be found on http://www.gleamviz.org and in Reference [3-5].

Real COVID-19 Data used for this study is available on following repository maintained by the Center for Systems Science and Engineering (CSSE) at Johns Hopkins University and “Our World in Data” website page “Coronavirus Pandemic (COVID-19)” https://github.com/CSSEGISandData/COVID-19 https://ourworldindata.org/coronavirus

## 3 Model compartmentalization

A modified SEIR metapopulation transmission model was adopted from [1] to simulate the spread of the COVID-19 epidemic in Pakistan. In this model, seven different population compartments are used: 1. Susceptible –total susceptible individuals. 2. Latent non-infectious 3. Presymptomatic infectious –Individuals who are infectious but show no symptoms so far 4. Mild symptoms – Individuals with mild and asymptomatic cases 5. Severe symptoms – Individuals with strong symptoms and are supposed to be quarantined. 6. Diagnosed – Individuals with confirmed diagnosis of COVID-19. 7. Recovered – The individuals who either got recovered or died (*Fig:1*).

**Figure 1:**
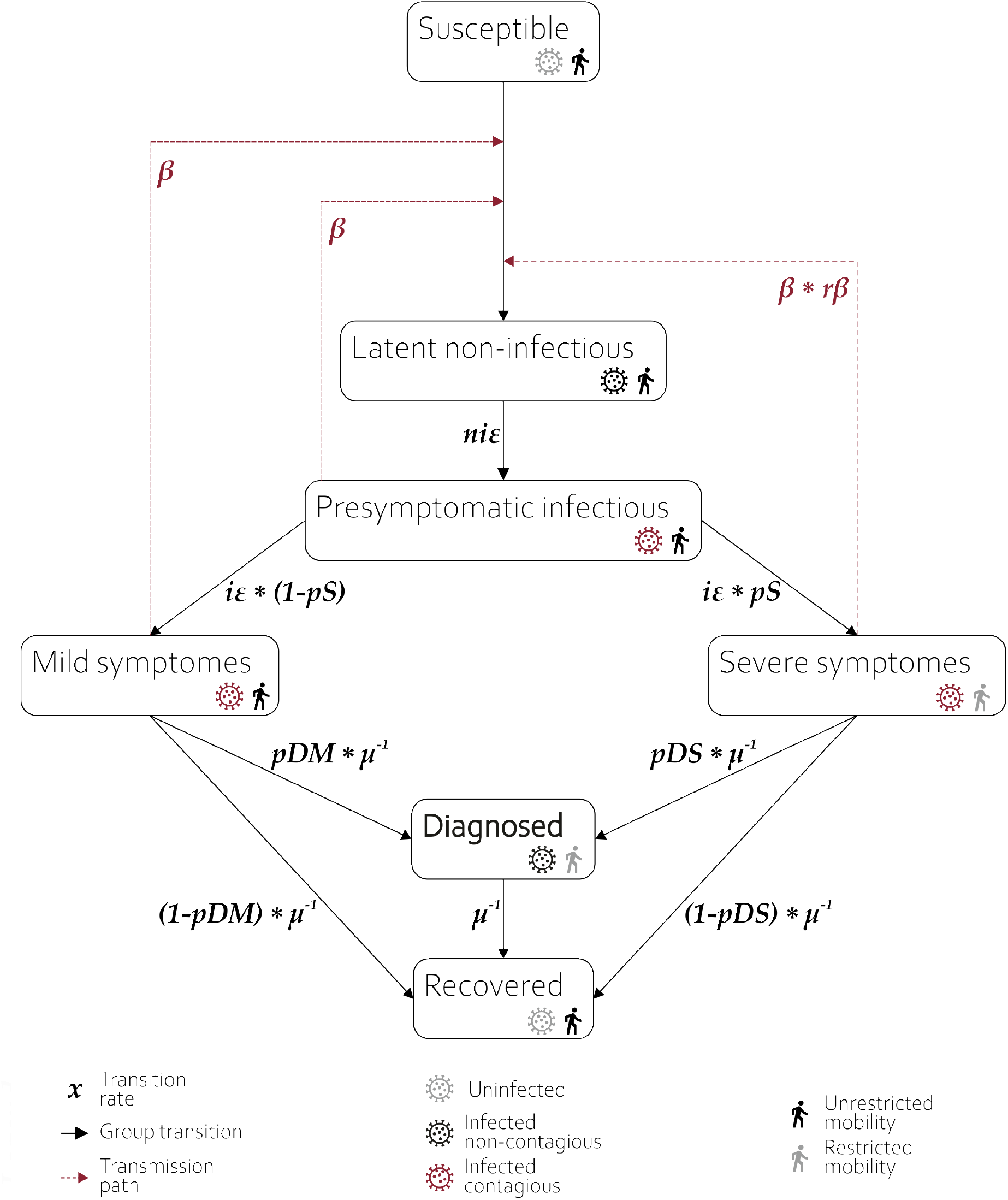
The structure of model and compartmentalization.

A simplified scheme for flow of model can be described as following.

A susceptible individual may get in contact with a person who is either: 1) presymptomatic with contact rate β 2) who developed mild symptoms with rate β, or 3) who developed severe symptoms and may contract the infection at rate *r*β*β. He then enters the latent non-infectious compartment where he is infected but not yet infectious. There he has a probability of *niε* of becoming presymptomatic infectious. The presymptomatic cases have chances i*ε* of developing severe symptoms with probability *pS* or mild symptoms, with probability (1-*pS)*. Individuals with severe symptoms are not allowed to travel within and between modelled subpopulations and may be either diagnosed with probability *pDS*, or recover with probability of 1-*pDS*. Individuals having mild (or non-existent) symptoms are allowed for traveling and they may be diagnosed with probability *pDM* (or recover with probability 1-*pDM)*. The diagnosed individuals are isolated and effectively non-contagious and they recover with rate *μ*. The recovery does not discriminate between true recovery and fatal cases.

## 4 Model parametrization

Since the accuracy of a simulation depends on the selection of its parameters, we used both, the relevant literature and educated estimation-based methods for making a choice for initial parameters.

We can divide our model parameters in two types:

### a) Literature based evident parameters

Literature based parameters are those that are fixed for this COVID-19 epidemic and likely do not change significantly across countries or regions or states. These parameters are, latency period ***lp***, probability for development of the severe condition ***pS***, and average recovery time after symptoms development **μ**. We used a value for lp as 5.6 days, pS 0.01 and **μ** as 7 days based on the discussion in [1].

However, latency period of COVID-19 appears to be sufficiently longer as compared to other similar viruses with pre-symptomatic transmission [8,9]. Therefore, it is plausible to divide the latency period in two sub-categories namely, average latent non-infectious period ***lnip*** and average presymptomatic infectious period **pip**. The values for these parameters are chosen as 1.1 days and 4.5 days respectively based on the studies performed in [1]. We therefore have two more parameters for the simulation, non-infectious latency rate - non-infectious epsilon ***niε*** and infectious latency rate - infectious epsilon ***iε***. These are specified as following

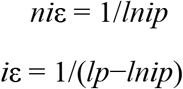

A more detailed discussion on these parameters can be found in [1].

### b) Local conditions based estimated parameters

The variable parameters are those that may vary for country, region, or state to state. For our model these parameters are:

Reproductive number, **R_0_:**

There is a number of published research papers available on the efforts made for estimation of R_0_ with values ranging from 1.4 to 6.49 [6]. However, WHO maintains a rather narrow range with 1.4– 2.5 and a study by Liu et al estimated an average R_0_ for COVID-19 to be 3.28 [10]. We decided to use R_0_ as 2.875(pre-lockdown) for Pakistan which lies well within the range of these studies.

Detection probability for a severe infection, ***pDS***:

Siwiak et.al [1] used a value of 0.6 for their studies, however since the condition of testing facilities and public response regarding clinical procedure in Pakistan are way less as compared to an “average” global value used for that study. We therefore, decided to use a value of 0.06 for our simulation. However, as the testing facilities improved by import of testing kits and public awareness to go for testing etc, we raised it gradually to 0.13.

Rate of total diagnosed to undiagnosed cases, **tDR:**

As the rate of diagnosed cases is suspected to be very less for developing countries like Pakistan, we adopted the formula for estimating the minimum value of tDR from [1]

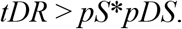

Detection probability for a mild infection, ***pDM:***

pDM can be derived from previously estimated parameters by using following equation [1]

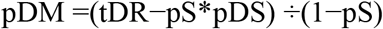

Effective contact rate, ***β:***

Effective contact rate β is derived using the following relations [1]

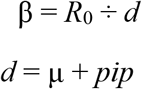

An associated parameter ***rβ***, the reduction level for patients who had the severe symptoms but were not diagnosed is set to 0.5 based on previous studies for COVID-19 and 2009 influenza outbreak [1]

## 6 Results and discussion

The simulation starts with a single host in Wuhan, China and the results of global spread based on mobility and transmission model gives reasonable agreement with the data for several countries across four continents within the 95% confidence band [1].

For Pakistan, we considered the several local factors that could introduce a deviation in the trend for COVID-19. For example, the partial/stepwise lockdown imposed by government from 25^th^ March till 09^th^ May, 2020. However, we noted that the effect of lockdown increased during Ramadan (24^th^ April – 09th May) as the legislators opted for a strategy of allowing only specific necessary stores of food/household items to remain open during 09 am – 05 pm and fasting season reduced the mobility even more. On 09^th^ May, authorities lifted the lockdown thereby allowing increased population mobility. Despite the restrictive instructions from government of Pakistan, due to Eid intense shopping and celebration preparations the coronavirus rate increased abruptly. From 1^st^ till 20^th^ June, we assumed a slightly reduced mobility compared to Eid-season, but higher than lockdown. And from 21^st^ June, we assumed the mobility to be back again with a very minute difference from the original value before lockdown. This reduced mobility is to take into consideration the social distancing measures that public is supposed to be following as compared to the time regime before 25, March 2020 (lockdown). After translating all these effects and a gradual increase in testing facilities during this period into the simulation parameters, we obtained a reasonable agreement in simulated and actual data.

Figs. 2-3 show the cumulative diagnosed and cumulative death cases in Pakistan. The chosen parameters R_0_ and μ that best describe the epidemic are 2.875 (without intervention) and 7.0, respectively. To estimate the death cases, we scaled down the diagnosed cases with an assumed average fatality rate of 2.1% as provided by Government of Pakistan [11]. The simulation agrees with the real data as of 1^st^ June, 2020. Figs. 4-5 show the daily diagnosed and death cases. We extended the simulation to estimate the peak time, maximum daily and cumulative cases. We found that under the current scenario, peak occurs in second decade of June, 2020 with (3600-4200) daily cases. The current wave of SARS-COV-2 is estimated to cause some (210,000 – 226,000) cumulative cases and (4400-4750) cumulative lost lives by the end of August in Pakistan.

**Figure 2:**
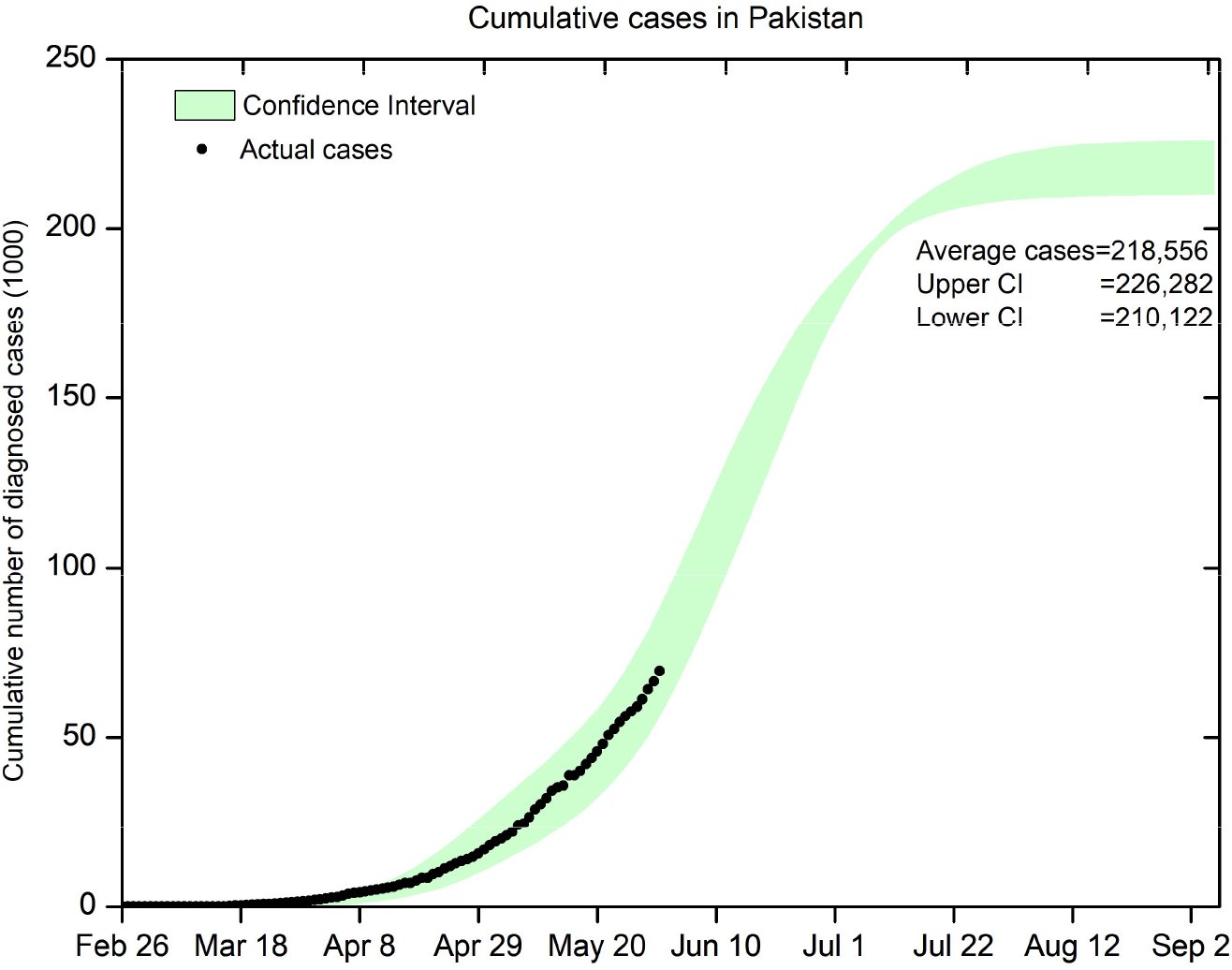
Cumulative Diagnosed cases in Pakistan.

**Figure 3:**
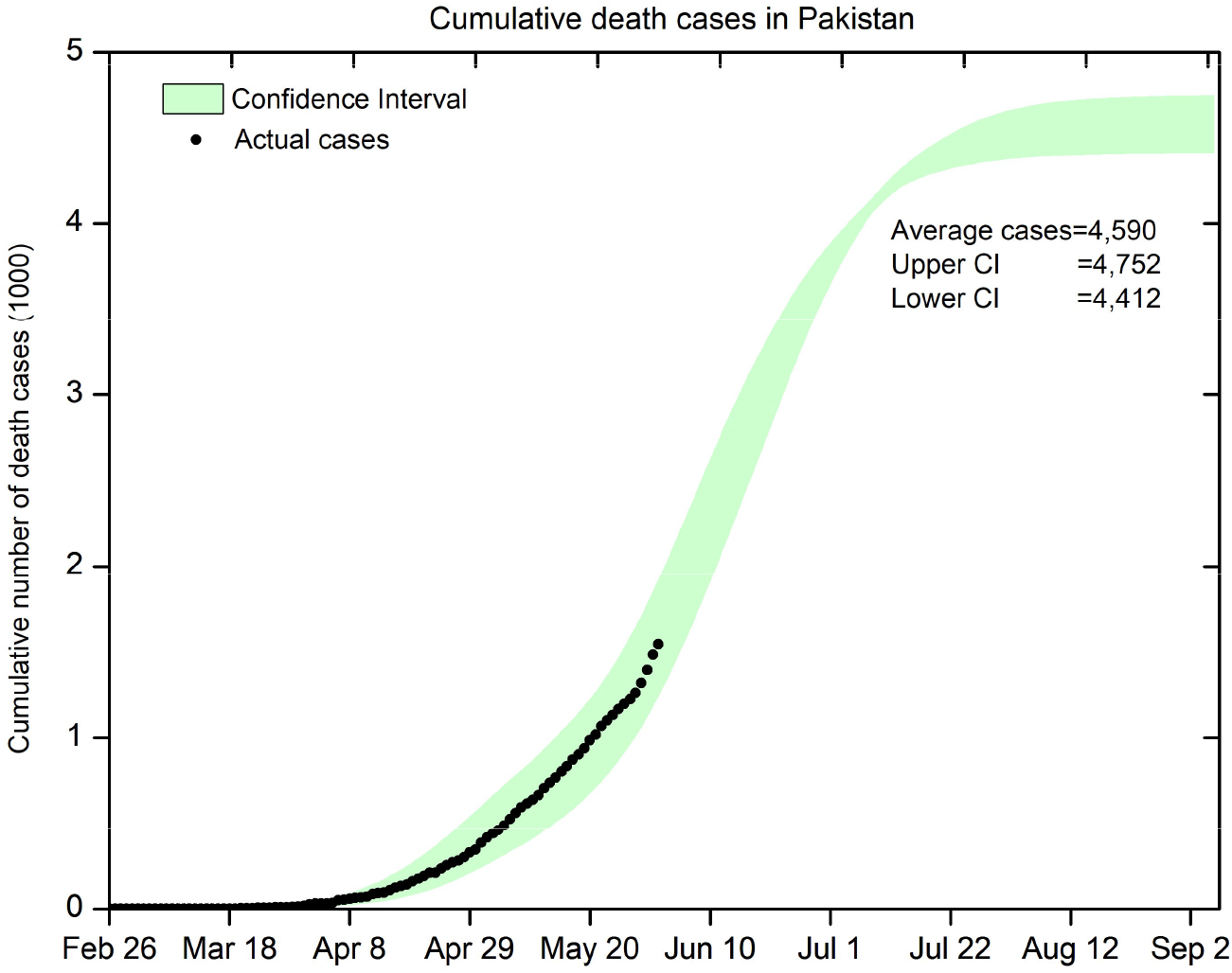
Cumulative death cases in Pakistan.

**Figure 4:**
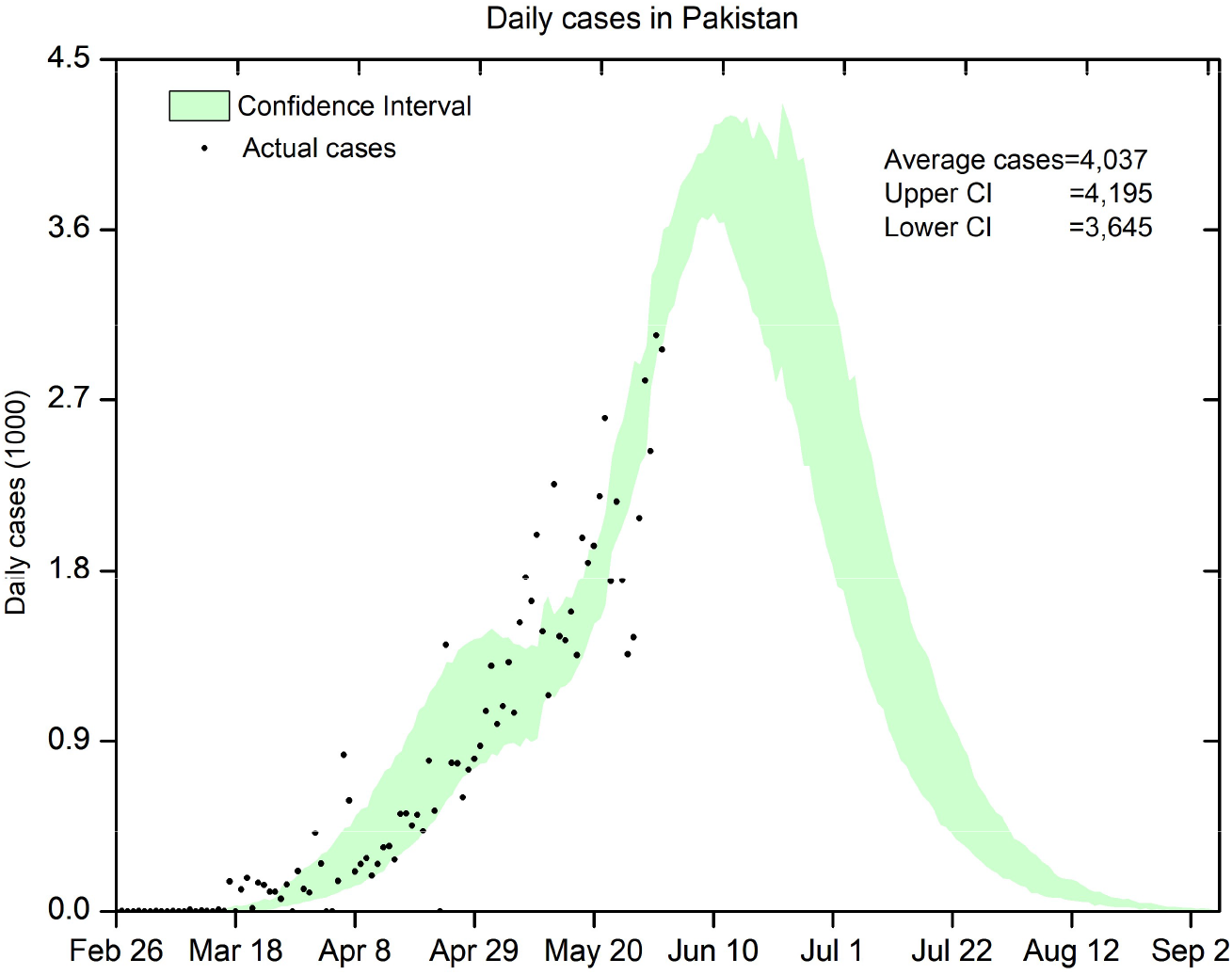
Daily Diagnosed cases in Pakistan.

**Figure 5:**
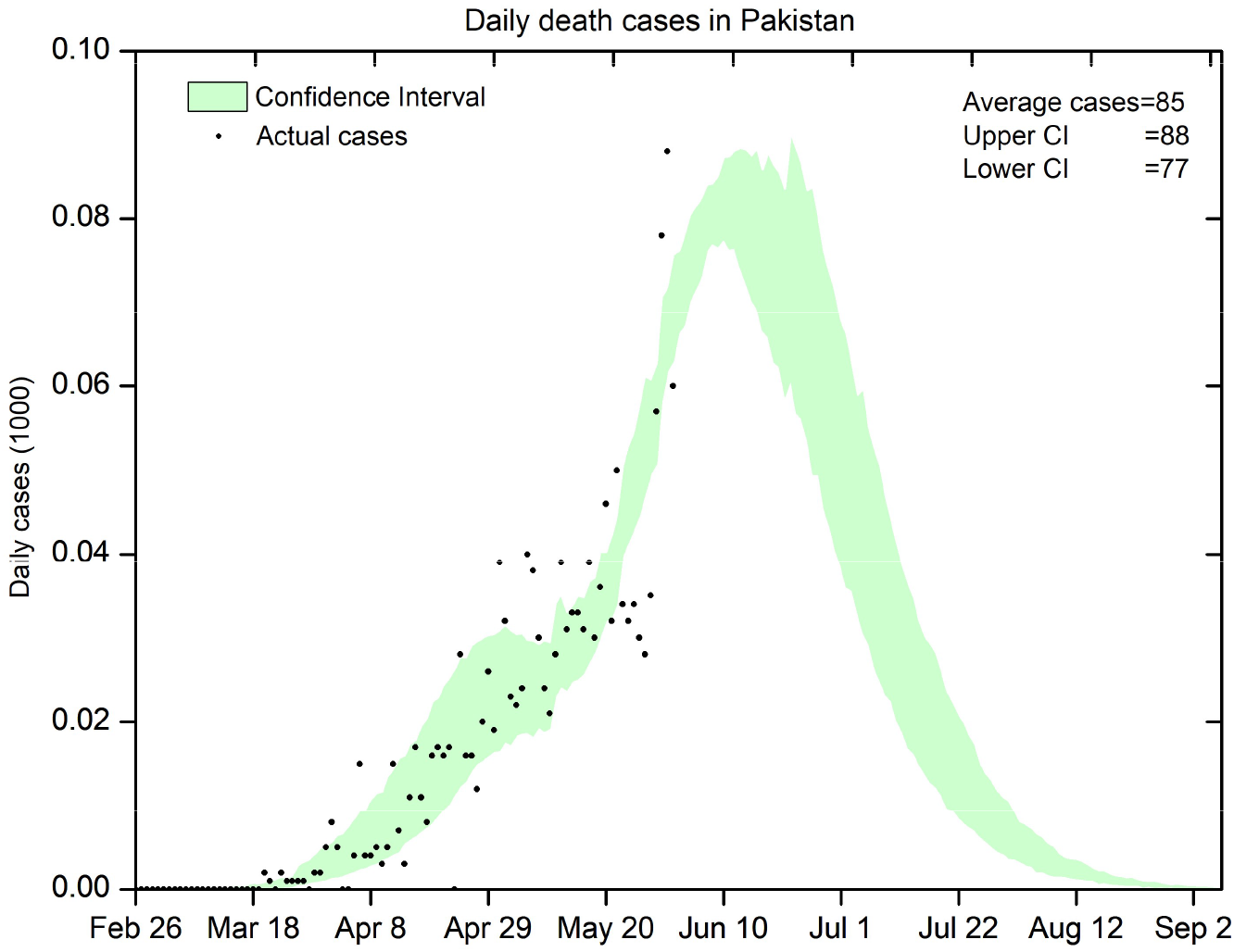
Daily death cases in Pakistan.

To obtain the best fitting we considered several variations of assumed parameters R_0_ and μ. Figs. 6-7 depict the effects of variation in effective reproductive number R_0_ (2.725, 2.775, 2.875, 2.975, 3.025) and the average recovery time μ (5, 7, 8, 9, 10), respectively, and indicate the selection of R_0_ value 2.875 and μ value as 7.0 best describe the real data.

**Figure 6:**
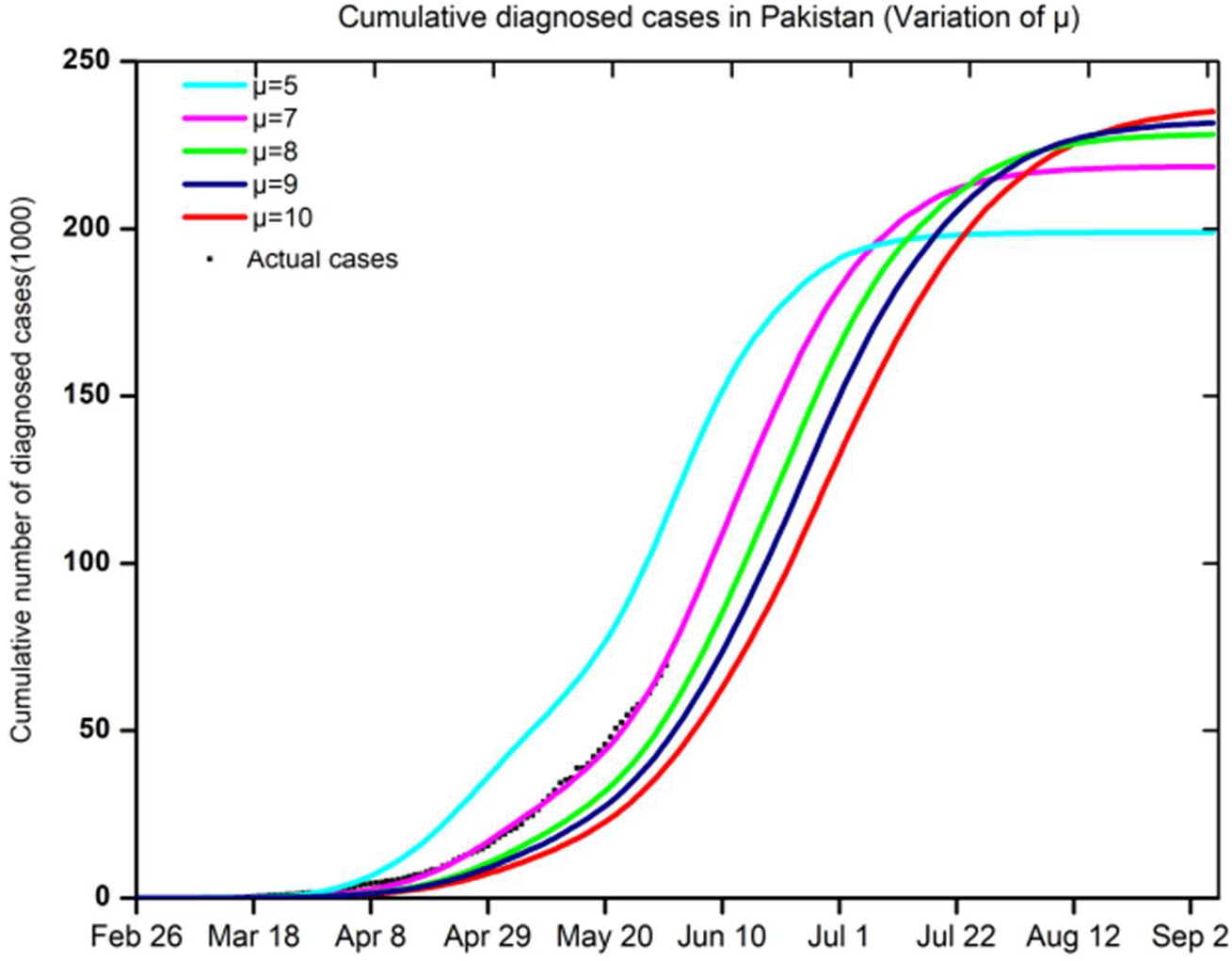
Variation of μ (average recovery time).

**Figure 7:**
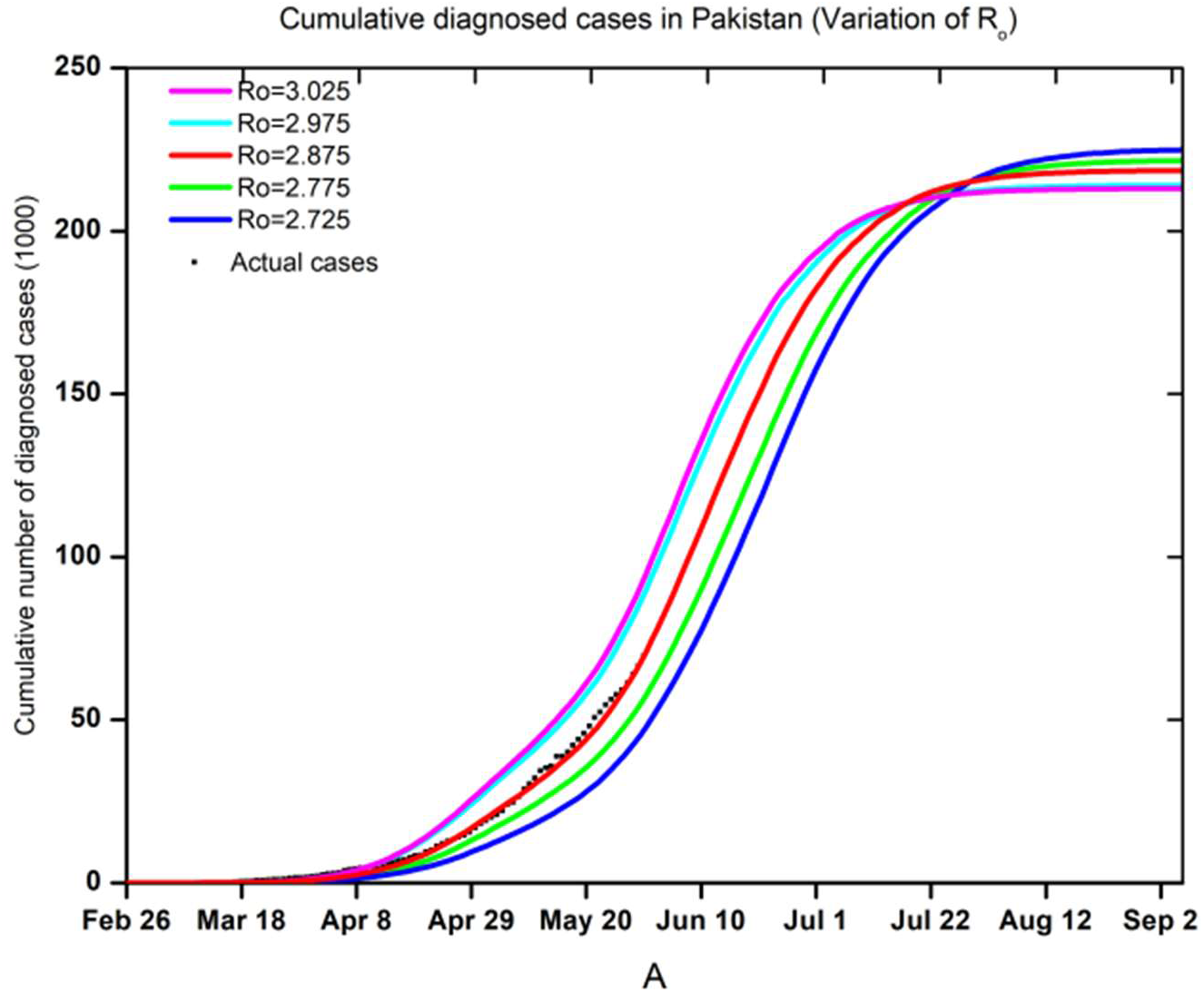
Variation of R_0_ (reproduction number).

To investigate the effects introduced by lockdown imposition on COVID-19 trend in Pakistan, we simulated four cases with corresponding parameters Figs. 8-11.

1. -No LockDown
2. -Partial/Step wise Lockdown (Actual Case)
3. -Strict LockDown (4 months)
4. -Actual Case with proposed Lockdown in June

**Figure 8:**
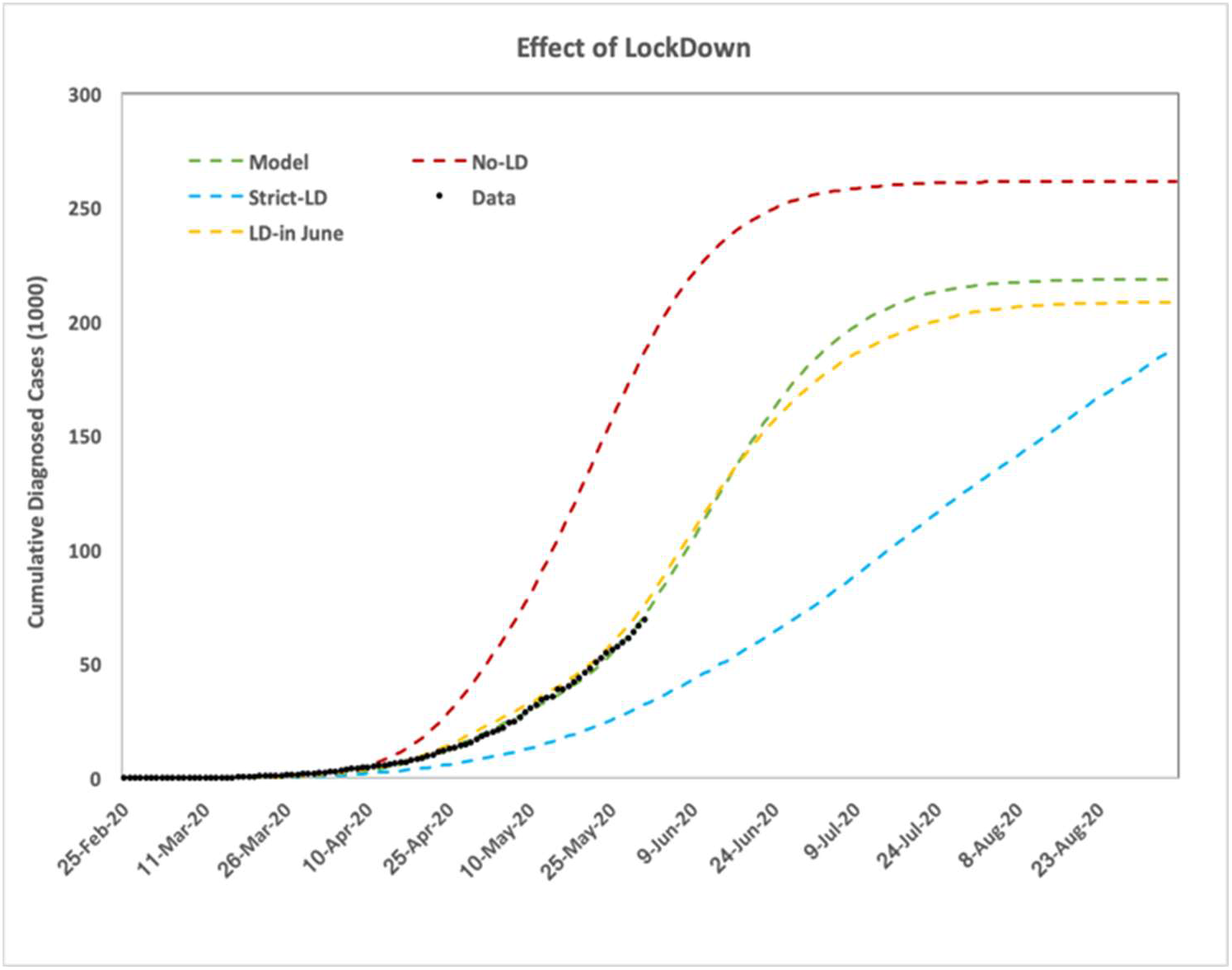
Lockdown impact (Cumulative Diagnosed Cases).

**Figure 9:**
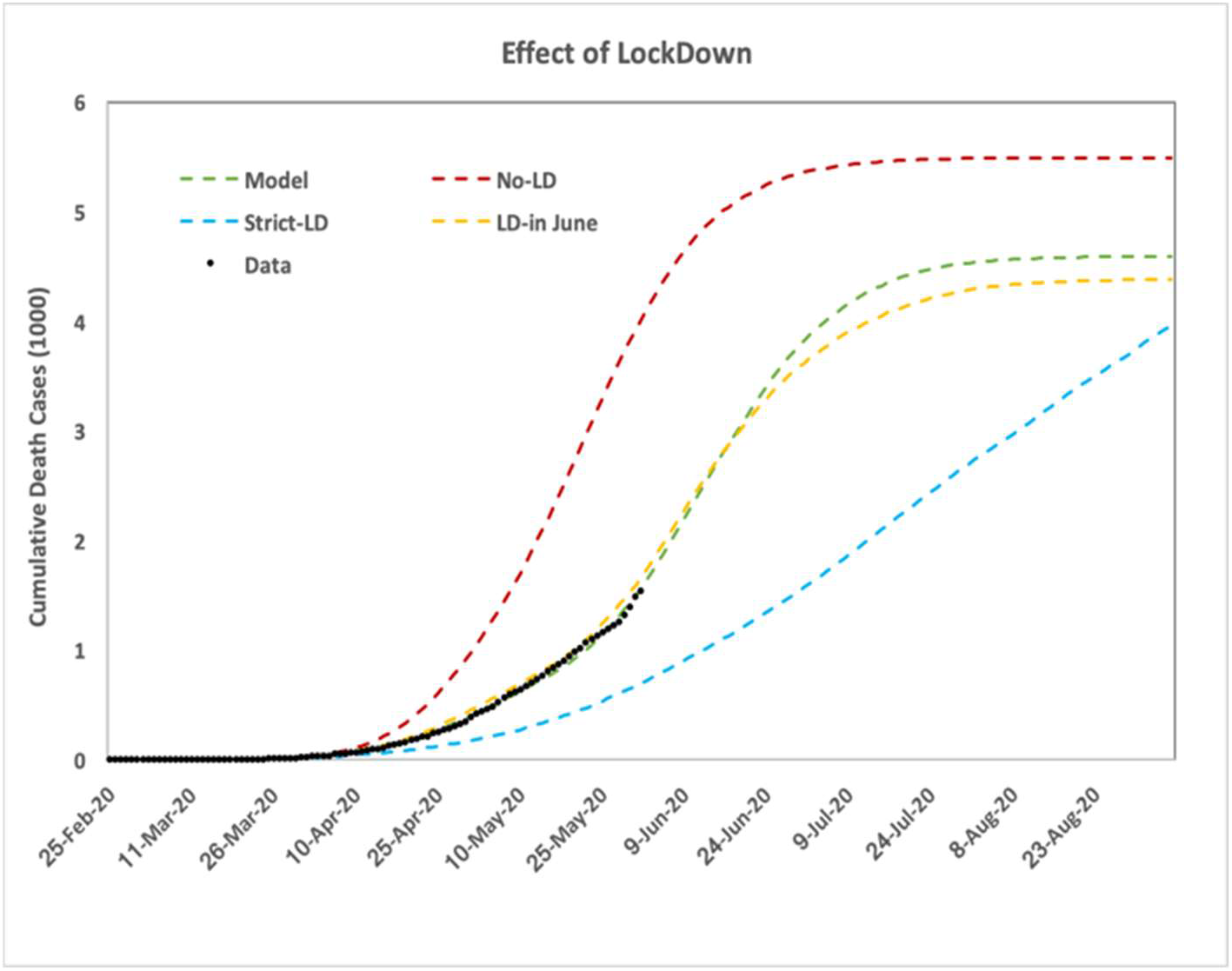
Lockdown impact (Cumulative Deaths).

**Figure 10:**
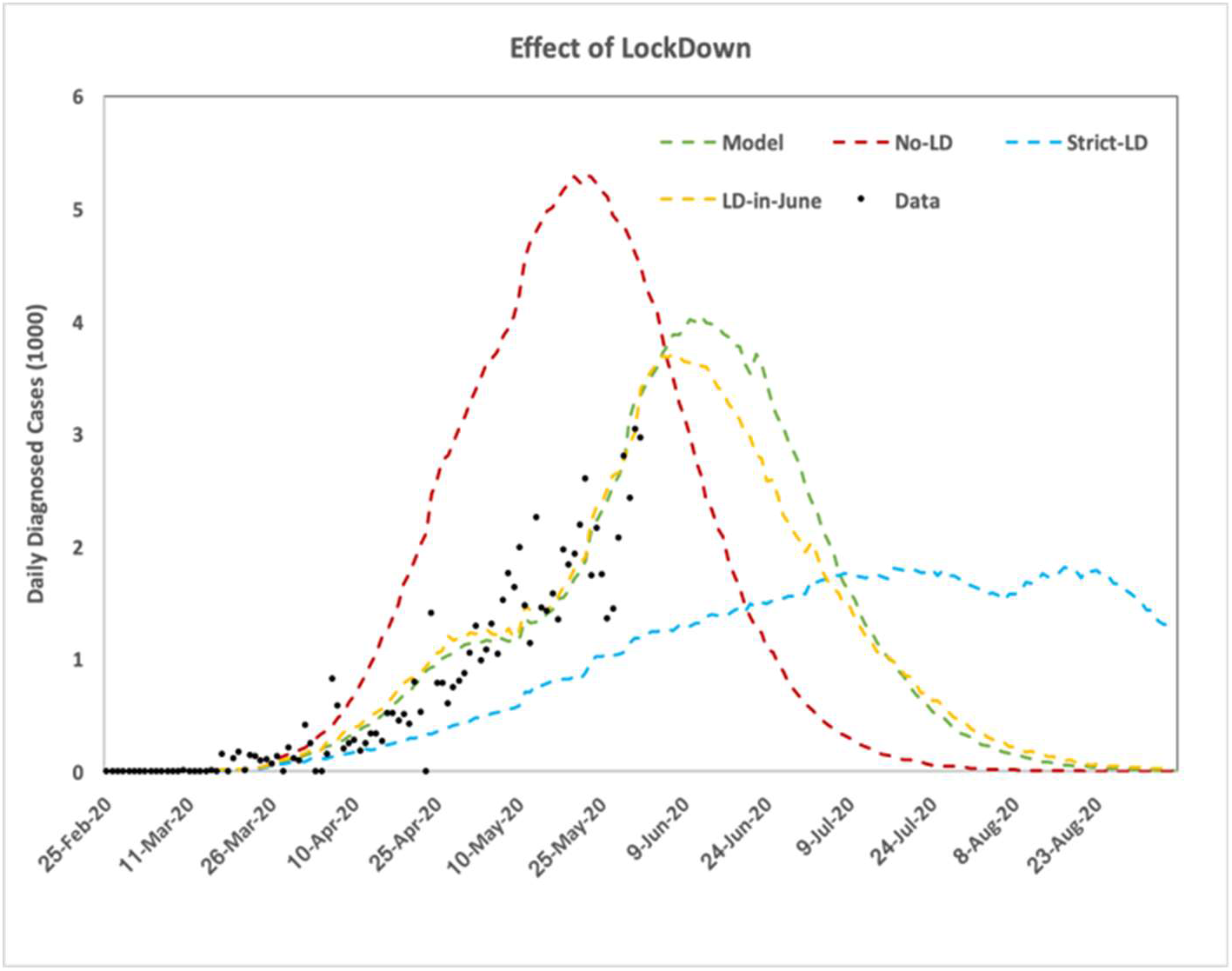
Lockdown impact (Daily Diagnosed Cases).

**Figure 11:**
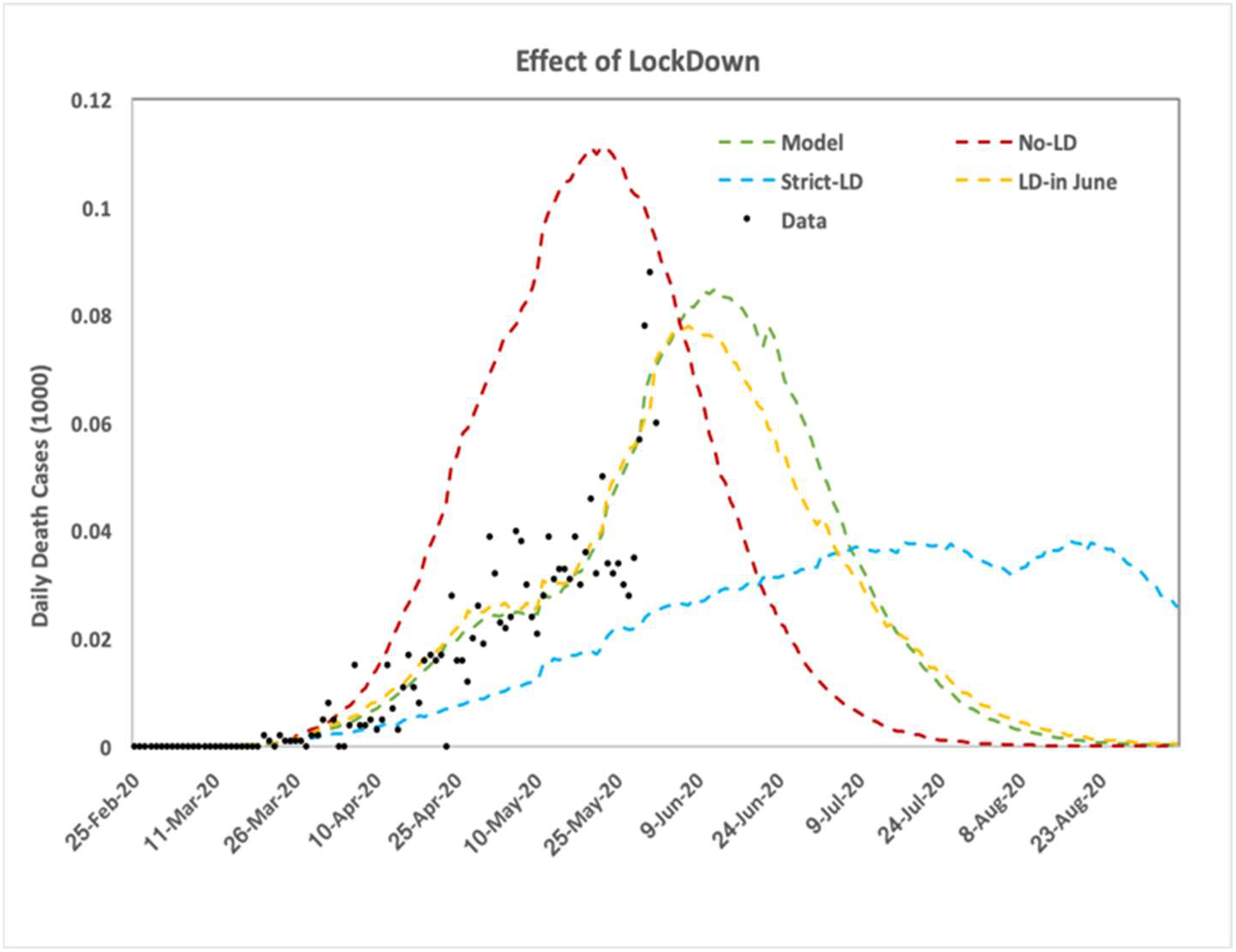
Lockdown impact (Daily Deaths).

The lockdown, social distancing measures and contact tracing and best strategies to contain an epidemic. The lockdown reduces the stress on hospital beds, number of health workers and testing capabilities. It buys time for a country to prepare its health necessities by shifting the peak in the future. Moreover, it reduces the cumulative number of infected population and hence saves lives in the long run. Pakistan imposed a lockdown (24^th^ March – 09^th^ May) and we also considered the fact that people mobility reduced even further during Ramadan (23^rd^ April-23^rd^ May) thereby increasing the effectiveness of lockdown. Despite the big questions on compliance with lockdown in Pakistan, Figs. 8-9 show that the cumulative diagnosed and death cases are reduced. We added a hypothetical case in which lockdown goes on for four months (25^th^ March-25^th^ July) and it further reduces the burden on health care and cumulative cases. But this strategy would severely damage the economic cycles. With current lockdown timeline we considered a possibility of imposing 2^nd^ phase lockdown in June, as was under consideration by authorities. This lockdown would yield better results in reducing the cumulative deaths/infections as well as in bending the curve, therefore putting less burden on the healthcare system. This type of lockdown is a recommended option in view of balancing the economic wheel to move on and containing the epidemic. However, as the Government of Pakistan announced the lifting of lockdown, it is extremely necessary to follow social distancing protocols at individual levels and keeping the epidemic in control during this peak duration. The following table summarizes the effects of different lockdown strategies considered in the study.

**Table 1:** Lockdown impact.

| Lockdown strategy | Max. Daily Diagnosed Cases | Max. Daily Death Cases | Cumulative Diagnosed Cases | Cumulative Death Cases | Peak Time | 95% Decay Time | 99% Decay Time |
| --- | --- | --- | --- | --- | --- | --- | --- |
| No-LD | 4250 - 5450 | 89 - 1128 | 256k – 265k | 5400 - 5570 | 20-27 May | 5-10 July | 20-27 July |
| Actual-LD | 3600 - 4200 | 77 - 89 | 210k – 226k | 4400 - 4700 | 7-17 June | 1-9 August | 17-27 August |
| Actual-LD+June-LD | 3400 - 3980 | 71 - 83 | 198k – 221k | 4175 - 4650 | 7-15 June | 2-18 August | 23Aug-5Sep |
| 4months-LD | 1655 - 1930 | 34 - 40 | 182k – 192k | 3822 - 4000 | 15-20 July | Mid- Sep | End-Sep |

## 7. Conclusions

Modeling and simulation have been performed using GLEAMviz for prediction of COVID-19 in Pakistan. It has been found that careful selection of parameters, based on the local conditions, could model the epidemic in Pakistan. The recent selection of parameters and epidemic prediction is based on the number of tests in Pakistan and the selection of parameter R_0_ as 2.875 as compared to 4.4 in [1] indicate that Pakistan is doing less tests per million as compared to other countries. Similarly, we have used the value of detection probability for a severe infection, *pDS* to be 0.06 and then gradually raised to 0.13, whereas it has been used 0.6 in [1]. With the increase in the number of tests per million the value of R_0_ and *pDS* may improve to better estimate the epidemic. With the recent selection of parameters, the peak of COVID-19 in Pakistan is expected around second decade of June while maximum number of cases and deaths could reach upto approximately (210k-226k) and (4400-4700) respectively. The epidemic is expected to last till September with 95% decay in first week of August and 99% decay in the last week. The Lockdown does have an impact of containing the epidemic in Pakistan and we see reduced cumulative and daily numbers. However, to use its impact effectively we should consider a smaller second phase lockdown in start of June or we must follow restrictive social distancing protocols individually.

## Data Availability

Data is not available online

## References

[1] Marlena M Siwiak et al., “From a single host to global spread. The global mobility based modelling of the COVID-19 pandemic implies higher infection and lower detection rates than current estimates,” medRxiv, 2020 May 2020.

[2] worldometer, “COVID-19 CORONAVIRUS PANDEMIC,” [Online]. Available: https://www.worldometers.info/coronavirus/.

[3] Wouter Van den Broeck et al., “The GLEaMviz computational tool, a publicly available software to explore realistic epidemic spreading scenarios at the global scale,” BMC Infectious Diseases, vol. 11, 02 February 2011.

[4] Duygu Balcan et al., “Seasonal transmission potential and activity peaks of the new influenza A(H1N1): a Monte Carlo likelihood analysis based on human mobility,” BMC Medicine, vol. 9, 10 September 2009.

[5] Duygu Balcan et al., “Modeling the Spatial Spread of Infectious Diseases: The GLobal Epidemic and Mobility Computational Model,” Journal of Computational Science, vol. 1, no. 3, pp. 132-145, August 2010.

[6] Michele Tizzoni et al., “Real-time numerical forecast of global epidemic spreading: case study of 2009 A/H1N1pdm,” BMC Medicine, vol. 10, 13 December 2012.

[7] Kashif Zia et al., “COVID-19 Outbreak in Pakistan: Model-Driven Impact Analysis and Guidelines,” arXiv.org, 31 March 2020.

[8] Roman Woelfel et al., “Clinical presentation and virological assessment of hospitalized cases of coronavirus disease 2019 in a travel-associated transmission cluster,” medRxiv, 2020 March 2020.

[9] Lauren Tindale et al., “Transmission interval estimates suggest pre-symptomatic spread of COVID-19,” medRxiv, 06 March 2020.

[10] Fatima Mukhtar, Neha Mukhtar, Jounal of Ayub Medical College Abbottabad, vol. 32, no. 1, pp. 141-4, 2020.

[11] Government of Pakistan, “Coronavirus in Pakistan,” [Online]. Available: http://covid.gov.pk/.

